# Post-Hurricane Fluid Conservation Measures Fail to Reduce IV Fluid Use in Critically Ill Children

**DOI:** 10.1101/2025.02.04.25321427

**Authors:** Celeste G. Dixon, James Odum, Ulka Kothari, Susan D. Martin, Julie C. Fitzgerald, Ami Shah, Heda Dapul, Chloe G. Braun, Andrew Barbera, Nina Terry, Scott L. Weiss, Denise C. Hasson, Adam C. Dziorny

## Abstract

**Introduction:** Fluid overload (FO) due to excessive fluid administration in critically ill children is associated with worse pediatric intensive care unit (PICU) outcomes. Damage to intravenous fluid (IVF) production plants due to Hurricane Helene resulted in a national IVF shortage, leading many hospitals to implement IVF conservation guidelines. We hypothesized that these guidelines would lead to a reduction in IVF use and a decrease in % cumulative fluid balance (CFB).

**Methods:** Four-site cohort study of critically ill children utilizing a federated data collection framework to extract patient age, sex, weight, and daily fluid intake/output for all admissions one month prior to and one month after the implementation of IVF conservation guidelines. Guidelines were individualized per institution. Total fluid intake, total IVF intake, % intake from IVF, and % CFB, as total intake minus total output divided by PICU admission weight, were compared between pre and post IVF conservation groups.

**Results:** There were 633 pre and 619 post IVF conservation encounters, with similar age and size distributions. There was no significant difference in IVF use pre and post IVF conservation. All sites had similar conservation recommendations.

**Conclusions:** Simple recommendations without structured change was insufficient to change IVF administration practices, indicating additional practices will be needed to reduce IVF intake and % CFB in this vulnerable population.

## INTRODUCTION

Critically ill children in the pediatric intensive care unit (PICU) frequently receive intravenous fluids (IVF) for resuscitation, maintenance of hydration, and medication delivery. While IVF administration is often necessary, there are risks associated with excessive fluid administration. Excessive positive cumulative fluid balance (CFB), is associated with higher mortality and longer PICU length of stay (LOS) (1–3). Due to these associations, there have been recent efforts to evaluate the amount of IVF prescribed to PICU patients and find opportunities for improvement (4).

On September 29^th^, 2024, Hurricane Helene caused catastrophic damage and flooding in western North Carolina, near the production plant of Baxter International (Deerfield, IL), a major producer of IVF. The resulting halt in production and distribution led to an acute national IVF shortage, stressing hospital systems and leading to a call for providers to more carefully consider IVF use in their patients (5).

Our existing multicenter collaborative has been studying ways to reduce CFB in children with critical illness. In response to this crisis, each of our academic PICUs implemented IVF conservation guidelines. The objective of this study was to identify associations between fluid conservation efforts and CFB. We conducted a multicenter retrospective cohort study of IVF administration practices one month prior to, and one month post, the initiation of IVF conservation initiatives. We hypothesized that hospital-wide mandates for IVF conservation would result in less IVF administered to PICU patients, and that we would observe variability among hospitals with different conservation policies.

## METHODS

### Study design

This is a retrospective observational study including patients admitted to the pediatric ICU at four tertiary care centers. Each sites’ institutional review boards (IRB) deemed this study exempt from IRB review. This study was conducted in accordance with the ethical standards of the Helsinki Declaration of 1975. All sites identified the date that IVF conservation strategies were implemented in their ICU (“implementation date”) and described their approaches to IVF conservation (**Figure 1**). We utilized a federated data collection framework to collect and aggregate clinical data without sharing row-level data among sites (6). We updated previously written data extraction queries and analysis scripts for this study.

**Fig. 1.**
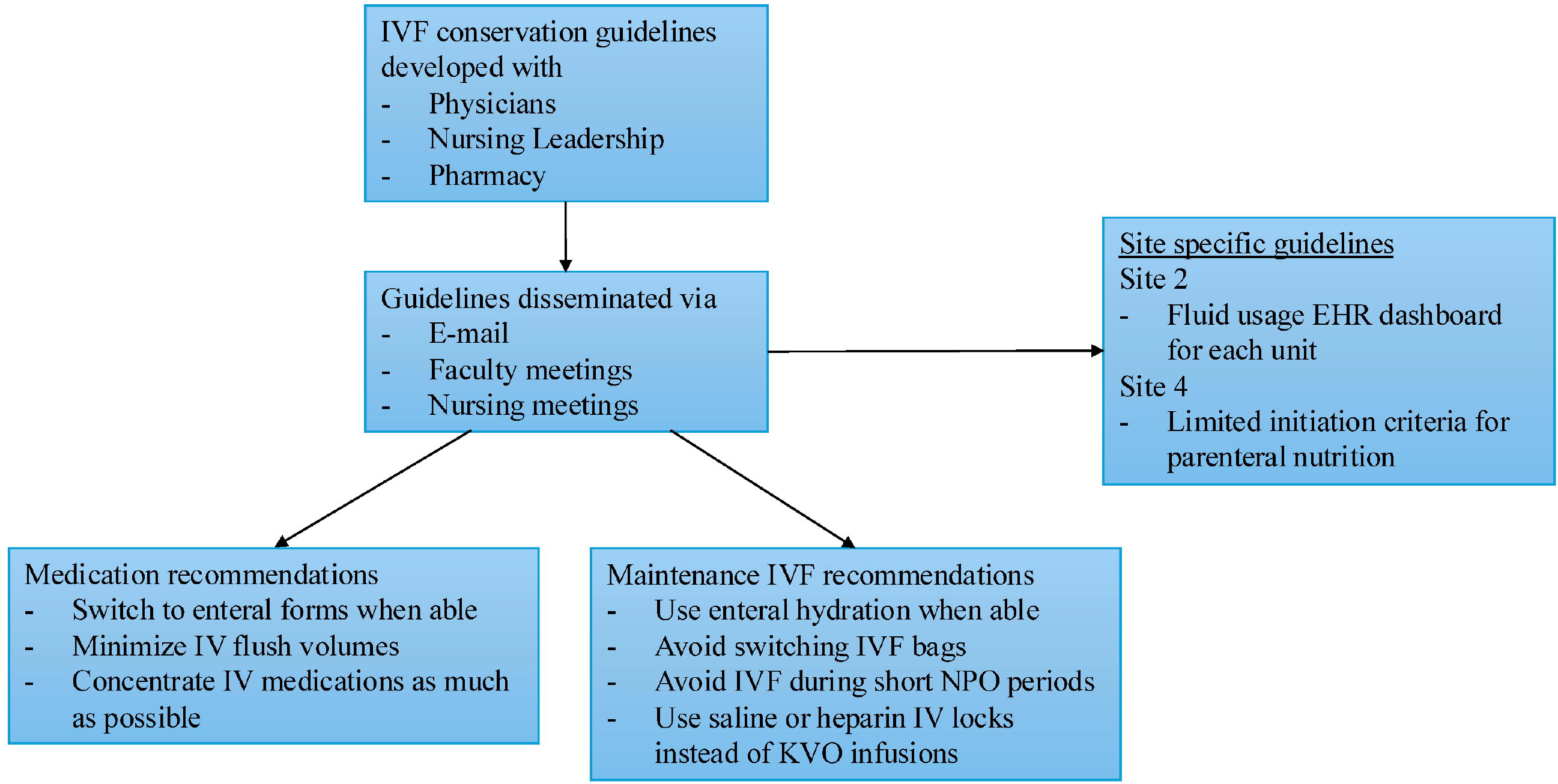

We compared all ICU admissions in the month prior to each site’s identified implementation date (“pre”) to the month following this date (“post”). We included a four-day washout period surrounding this date. For each ICU encounter, we recorded patient demographics (age, admission weight), fluid intake (amount, route), urine output and occurrences (output that occurred but could not be quantified), and LOS from the electronic health record (EHR). Each site extracted their own data and ran identical analysis scripts.

### Analysis

Consistent with prior studies (1), we defined Day 0 of ICU stay as admission time through 7 AM the following day. We defined Day 1 as the subsequent 24 hours (7 AM to 7 AM), and Day 2 as the following 24 hours. Total intake and IVF intake were standardized to ml/kg/hr. We calculated fluid metrics for Days 1 and 2, including: Percent of total intake as IVF as *total IVF intake/(total enteral + IVF intake)*; Percent of Holliday-Segar received as *total IVF/(4x first 10 kg weight + 2x number of kg between 10-20 + 1x number of kg >20)* (7); and Percent CFB as *[cumulative fluid in (L) – cumulative fluid out (L)]/PICU admission weight (kg) x 100* (8). We excluded ICU encounters with urine occurrence counts from CFB analyses so as to not exaggerate CFB values. Differences between groups were compared using Chi-square test of independence, Fisher’s exact test, or Wilcoxon rank sum as appropriate using analysis scripts written in R (https://www.r-project.org). P values of <0.05 were considered significant.

## RESULTS

### Fluid measurements pre and post conservation

Cohort demographics are shown in **Table 1**. There were 633 pre encounters and 619 post encounters, with Sites 1 and 3 having more patients post and Sites 2 and 4 having fewer. Most sites had a slight male predominance, with Site 3 and 4 demonstrating a significant association between sex and temporal grouping. All sites had more younger (<1 year) and smaller (<10 kg) patients in the post group, although these differences were not significant. There was no significant difference in the number of patients with LOS >7 days at any site.

**Table 1:** Cohort and Fluid Metrics.

|  | Site 1 |  | Site 2 |  | Site 3 |  | Site 4 |  |
| --- | --- | --- | --- | --- | --- | --- | --- | --- |
|  | Pre | Post | Pre | Post | Pre | Post | Pre | Post |
| ICU Admissions (n) | 80 | 104 | 338 | 300 | 75 | 88 | 140 | 127 |
| Female (%) | 45 (56%) | 50 (48%) | 140 (41%) | 130 (43%) | **40 (53%) \*** | **33 (38%) \*** | **72 (51%) \*** | **49 (39%) \*** |
| Age (n, %) |  |  |  |  |  |  |  |  |
| <1 yr | 13 (16%) | 29 (28%) | 75 (22%) | 81 (27%) | 11 (15%) | 12 (14%) | 21 (15%) | 30 (24%) |
| ≥ 1 and < 4 | 19 (24%) | 25 (24%) | 82 (24%) | 67 (22%) | 23 (31%) | 35 (40%) | 35 (25%) | 28 (22%) |
| ≥ 4 and < 12 yrs | 22 (28%) | 31 (30%) | 97 (29%) | 88 (29%) | 24 (32%) | 25 (28%) | 40 (29%) | 26 (21%) |
| ≥ 12 yrs | 26 (33%) | 19 (18%) | 84 (25%) | 64 (21%) | 17 (23%) | 16 (18%) | 44 (31%) | 43 (34%) |
| Weight (kg) |  |  |  |  |  |  |  |  |
| <10 kg | 14 (18%) | 29 (28%) | 85 (25%) | 84 (28%) | 10 (13%) | 17 (19%) | 28 (20%) | 34 (27%) |
| 10-30 kg | 34 (43%) | 44 (42%) | 143 (42%) | 129 (43%) | 40 (53%) | 47 (53%) | 51 (36%) | 40 (32%) |
| ≥30 kg | 32 (40%) | 31 (30%) | 110 (33%) | 87 (29%) | 25 (33%) | 24 (27%) | 61 (44%) | 53 (42%) |
| Patients LOS >7 days (n, %) | 15 (19%) | 20 (19%) | 51 (15%) | 46 (15%) | 12 (16%) | 10 (11%) | 11 (8%) | 11 (9%) |
| **Fluid Measurements** |  |  |  |  |  |  |  |  |
| Total intake (IV + enteral) (ml/kg/hr) <sup>a</sup> |  |  |  |  |  |  |  |  |
| ICU day 1 | 2.45<br>[1.48, 4.25] | 3.27<br>[2.26, 4.28] | 3.34<br>[2.11, 4.44] | 3.30<br>[1.97, 4.65] | 3.34<br>[2.14, 4.26] | 3.60<br>[2.53, 4.28] | 2.69<br>[1.35, 4.38] | 2.82<br>[1.28, 5.01] |
| ICU day 2 | 2.72<br>[1.24, 4.72] | 3.35<br>[2.22, 4.30] | 3.02<br>[1.97, 4.45] | 3.21<br>[1.74, 5.27] | 3.25<br>[2.21, 4.30] | 3.37<br>[1.89, 4.33] | **2.65**<br>**[1.62, 4.09] \*** | **3.51**<br>**[2.02, 5.54] \*** |
| Total IV intake (ml/kg/hr) <sup>a</sup> |  |  |  |  |  |  |  |  |
| ICU day 1 | 1.56<br>[0.63, 3.30] | 1.44<br>[0.09, 3.26] | 1.74<br>[0.76, 2.82] | 1.50<br>[0.28, 3.18] | 2.75<br>[1.19, 3.91] | 2.13<br>[0.66, 3.70] | 1.36<br>[0.37, 2.52] | 0.94<br>[0.12, 2.97] |
| ICU day 2 | 0.96<br>[0.24, 2.83] | 1.44<br>[0.00, 3.16] | 1.00<br>[0.08, 2.45] | 1.02<br>[0.03, 2.65] | 1.56<br>[0.61, 3.38] | 1.11<br>[0.02, 3.34] | 1.36<br>[0.15, 2.58] | 1.01<br>[0.02, 2.76] |
| % of total intake as IVF <sup>a</sup> |  |  |  |  |  |  |  |  |
| ICU day 1 | **89.2**<br>**[45.5, 100.0] \*** | **64.9**<br>**[6.3, 100.0] \*** | 59.4<br>[25.7, 97.0] | 53.3<br>[15.1, 96.6] | 93.1<br>[64.0, 100.0] <sup>†</sup> | 76.9<br>[29.4, 100.0] <sup>†</sup> | 70.6<br>[26.7, 100.0] <sup>†</sup> | 58.9<br>[9.5, 95.4] <sup>†</sup> |
| ICU day 2 | 70.4<br>[17.2, 95.7] | 62.0<br>[0.0, 100.0] | 32.4<br>[4.1, 84.3] | 32.0<br>[3.3, 80.1] | 71.8<br>[17.7, 98.7] <sup>†</sup> | 42.1<br>[2.54, 85.8] <sup>†</sup> | 59.2<br>[10.9, 93.1] <sup>†</sup> | 40.5<br>[0.65, 80.2] <sup>†</sup> |
| % of calculated Holliday-Segar received <sup>a</sup> |  |  |  |  |  |  |  |  |
| ICU day 1 | 76.3<br>[36.4, 109.0] | 82.8<br>[41.6, 113.2] | 68.6<br>[34.5, 102.8] | 68.7<br>[25.5, 108.5] | 96.8<br>[50.2, 146.1] | 93.1<br>[46.3, 127.4] | 64.4<br>[23.8, 107.7] | 55.5<br>[18.0, 122.6] |
| ICU day 2 | 69.7<br>[25.3, 91.9] | 86.7<br>[41.0, 107.2] | 56.9<br>[18.4, 103.7] | 57.4<br>[14.7, 106.9] | 90.4<br>[31.7, 123.9] | 88.1<br>[28.8, 148.0] | 68.0<br>[27.8, 99.0] | 58.0<br>[20.9, 136.5] |
| >5% fluid overload <sup>b</sup> |  |  |  |  |  |  |  |  |
| ICU day 1 | 11 (30%) | 14 (29%) | 78 (31%) | 74 (34%) | 20 (35%) | 25 (39%) | 32 (29%) | 23 (27%) |
| ICU day 2 | 5 (33%) | 13 (52%) | 82 (45%) | 67 (42%) | 19 (54%) | 22 (48%) | 24 (39%) | 25 (50%) |
| >10% fluid overload<sup>b</sup> |  |  |  |  |  |  |  |  |
| ICU day 1 | 2 (5%) | 1 (2%) | 16 (6%) | 21 (10%) | 3 (5%) | 8 (13%) | 6 (6%) | 8 (9%) |
| ICU day 2 | 5 (33%) | 5 (20%) | 26 (14%) | 31 (20%) | 6 (17%) | 6 (13%) | 9 (15%) | 11 (22%) |

a) Patients with 0 ml intake excluded from this analysis.
b) Patients with urine occurrences (volume not recorded) excluded from this analysis.
<sup>†</sup>) Significant for $p \leq 0.1$
<sup>\*</sup>) Significant for $p \leq 0.05$

Day 1 and 2 fluid metrics are also shown in **Table 1**. Overall, IVF volumes on Days 1 and 2 of ICU admission were not consistently different between pre and post groups at any site. Total intake was either the same or slightly higher in the post group at all sites on both days, though this only reached significance at Site 4 on Day 2. IVF intake was lower in the post group at most sites; however, this difference was also not significant. The precent of total intake as IVF was variable across sites and decreased from pre to post. Although this decrease was only significant on Day 1 at Site 1 (p = 0.04) (Figure 1), Sites 3 and 4 showed a similar but nonsignificant trend on both Days 1 and 2 (*p* < 0.1). There was no significant difference in the number of patients with 5% or 10% fluid overload at any site on either Day 1 or 2. Across all sites, 29-35% of pre patients and 27-39% of post patients had ≥5% CFB on Day 1, with higher percentages of patients on Day 2 (33-54% pre and 42-52% post). A smaller percentage of patients reached ≥10% CFB, with a similar increase from Day 1 to Day 2 in both pre and post groups.

In supplementary analysis, the same metrics were compared for Day 3 and Day 4. Sites 3 and 4 had a significant decrease in percent of total intake as IVF on Day 4. No significant differences were observed for any other metric (**Supplementary Table 2**).

### Fluid conservation recommendations

All sites had similar recommendations to clinicians regarding IVF conservation practices (**Figure 1)**. Guidelines were created with input from physician, nursing, and pharmacy staff, and materials disseminated via e-mail and in-person meetings. Common themes identified were using enteral hydration when possible, converting medications to enteral forms, limiting use of fluids for short *nil per os* (NPO) times, and avoiding switching bags of fluid once started. No site required specific practice changes or utilized interruptive EHR alerts.

## DISCUSSION

In this four-site study of fluid practices surrounding Hurricane Helene, there was no significant difference in IVF administration or CFB between pre and post fluid conservation implementation. Despite nation- and hospital-wide mandates and the development of targeted guidelines, there was no observed change in clinical practice. It is not surprising that we also found no difference in the prevalence of positive CFB in the post groups.

These findings were consistent across PICUs in different geographic locations and of different sizes, and overall CFB >5% and >10% on Days 1 and 2 were very similar to percentages reported at the same sites over 6-month periods just 1.5 years prior (9). It is worth noting that some sites had a decrease in the percent of total intake given as IVF, while maintaining the same volume of total intake. This implies that rather than a reduction in IVF use, there was an increase in enteral fluid administration. There was a trend toward decreased IVF intake at Sites 3 and 4, although this was not significant.

It is likely that the reason for these null findings is not because changes were not attempted, but rather that more significant interventions are required to reduce CFB in critically ill children. Practice guidelines and recommendations related to passive fluid restriction, even within the context of a national IVF shortage, were insufficient to change amount of IVF administered. However, other groups have shown in single center prospective quality improvement studies that improved recognition of positive CFB and mandated decrease in maintenance IVF rates can reduce CFB (4, 10, 11). Although it is possible that targeting the de-resuscitation phase of illness may reduce IVF intake, we did not find differences when we extended our analyses to Days 3 or 4 (12).

There are limitations to this study. We did not collect data on type of fluid prescribed (e.g., balanced vs unbalanced crystalloids), why they were prescribed (maintenance vs carriers vs medications), or frequency of fluid bag changes. All sites did have recommendations to limit changing fluid type prior to finishing the bag, which may have reduced overall hospital IVF usage but not individual patient exposure. We also did not collect severity of illness measures, so it is possible that a discrepancy between pre and post groups could have accounted for some of the observed fluid administration. We believe this to be less likely given ICU LOS did not differ, and the CFB we observed was similar not only pre and post, but also to historical data from the same sites.

## CONCLUSION

Despite a national IVF shortage and specific IVF conservation guidelines, no significant difference in IVF use was observed during this time period. Other methods to mitigate IVF use and limit CFB should be investigated.

## Data Availability

All data produced in the present study are available upon reasonable request to the authors

