## Supplementary Data for "Post-Hurricane Fluid Conservation Measures Fail to Reduce IV Fluid Use in Critically Ill Children"

**Supplementary Table 1: Patient Counts by ICU Day**

|  | **Site 1** | | **Site 2** | | **Site 3** | | **Site 4** | |
| --- | --- | --- | --- | --- | --- | --- | --- | --- |
|  | **Pre** | **Post** | **Pre** | **Post** | **Pre** | **Post** | **Pre** | **Post** |
| Day 1 with urine occurrences | 72 | 96 | 334 | 299 | 69 | 85 | 139 | 133 |
| without urine occurrences | 63 | 88 | 331 | 299 | 66 | 82 | 136 | 126 |
| Day 2 with urine occurrences | 48 | 55 | 216 | 185 | 44 | 56 | 75 | 68 |
| without urine occurrences | 37 | 49 | 213 | 183 | 41 | 56 | 75 | 63 |
| Day 3 with urine occurrences | 35 | 41 | 148 | 133 | 31 | 33 | 49 | 37 |
| without urine occurrences | 27 | 39 | 147 | 132 | 31 | 32 | 49 | 37 |
| Day 4 with urine occurrences | 25 | 33 | 104 | 92 | 21 | 23 | 34 | 25 |
| without urine occurrences | 24 | 29 | 104 | 89 | 20 | 23 | 34 | 25 |

**Supplementary Table 2: Day 3 and 4 Fluid Metrics**

|  | **Site 1 (URMC)** | | **Site 2 (CHOP)** | | **Site 3 (NYU)** | | **Site 4 (UAB)** | |
| --- | --- | --- | --- | --- | --- | --- | --- | --- |
| **Fluid Measurements** | **Pre** | **Post** | **Pre** | **Post** | **Pre** | **Post** | **Pre** | **Post** |
| Total intake (IV + enteral) (ml/kg/hr) ^a^ | | | | | | | | |
| ICU Day 3 | 2.68  [0.72, 4.38] ^†^ | 3.58  [2.31, 5.28] ^†^ | 2.96  [2.07, 4.62] | 3.00  [1.66, 4.86] | 2.88  [1.96, 4.54] | 3.25  [2.14, 4.34] | **2.57**  **[1.65, 4.18]*** | **3.78**  **[2.04, 5.64]*** |
| ICU Day 4 | 3.02  [1.56, 4.57] | 3.44  [2.12, 5.23] | 3.53  [2.28, 5.39] | 3.67  [2.43, 5.84] | 3.10  [2.13, 3.82] | 3.38  [2.03, 4.93] | 2.96  [1.79, 5.05] | 4.17  [2.36, 6.19] |
| Total IV intake (ml/kg/hr) ^a^ | | | | | | | | |
| ICU Day 3 | 0.19  [0.00, 2.31] | 0.51  [0.00, 2.37] | 0.77  [0.00, 2.55] | 0.70  [0.00, 2.67] | 1.07  [0.23, 3.73] | 0.22  [0.00, 2.11] | 1.04  [0.10, 2.65] | 0.95  [0.15, 2.04] |
| ICU Day 4 | 0.65  [0.00, 2.50] | 0.36  [0.00, 1.85] | 0.56  [0.00, 2.24] | 0.85  [0.05, 3.23] | **2.49**  **[0.62, 3.30]*** | **0.53**  **[0.00, 2.06]*** | 0.96  [0.14, 2.57]^†^ | 0.29  [0.03, 1.04]^†^ |
| % of total intake as IVF ^a^ | | | | | | | | |
| ICU Day 3 | 43.3  [0.0, 100.0] | 42.0  [0.0, 88.5] | 19.7  [0.2, 86.1] | 20.7  [0.0, 68.8] | **68.2**  **[4.0, 99.1]*** | **18.0**  **[0.0, 66.2]*** | 64.0  [3.7, 95.0] | 25.4  [5.6, 77.5] |
| ICU Day 4 | 24.0  [0.0, 100.0] | 14.6  [0.0, 73.8] | 14.5  [0.0, 73.5] | 24.8  [2.0, 76.5] | **88.3**  **[33.9, 100.0]*** | **19.0**  **[0.0, 50.3]*** | **58.7**  **[9.8, 97.1]*** | **6.4**  **[0.8, 52.0]*** |
| % of calculated Holliday-Segar received ^a^ | | | | | | | | |
| ICU Day 3 | 15.1  [0.0, 75.9] | 26.7  [0.0, 86.1] | 24.3  [0.0, 97.1] | 22.7  [0.0,86.2] | 56.4  [7.5, 116.6] | 11.0  [0.0, 96.9] | 52.3  [3.8, 101.1] | 35.4  [5.7, 76.2] |
| ICU Day 4 | 16.2  [0.0, 84.5] | 19.9  [0.0, 76.8] | 19.6  [0.0, 83.1] | 25.7  [1.6, 94.3] | 77.7  [27.1, 136.3]^†^ | 19.3  [0.0, 129.3]^†^ | 41.7  [6.7, 121.4]^†^ | 10.3  [0.9, 33.1]^†^ |
| >5% fluid overload ^b^ | | | | | | | | |
| ICU Day 3 | 4 (40%) | 7 (58%) | 63 (57%) | 51 (53%) | 15 (65%) | 13 (62%) | 17 (43%) | 19 (63%) |
| ICU Day 4 | 3 (50%) | 8 (80%) | 44 (55%) | 42 (61%) | 10 (59%) | 7 (54%) | 15 (48%) | 16 (64%) |
| >10% fluid overload ^b^ | | | | | | | | |
| ICU Day 3 | 3 (30%) | 4 (33%) | 28 (25%) | 34 (35%) | 6 (26%) | 6 (29%) | **8 (20%)*** | **14 (47%)*** |
| ICU Day 4 | 3 (50%) | 4 (40%) | 33 (41%) | 28 (41%) | 3 (18%) | 4 (31%) | 10 (32%) | 12 (48%) |

a) Filtered for patients who had > 0 ml total fluids for given day.

b) Patients with urine occurrences (volume not recorded) excluded from this analysis.

^†^) Significant for *p* ≤ 0.1

*) Significant for *p* ≤ 0.05

**Fluid conservation recommendations by site**

Site 1: Notice of anticipated fluid shortage was distributed via internal communication on 10/2/2024. The following strategies were recommended: utilize any IV fluids in each bag before changing, including changes in fluid type; re-evaluate the need for ongoing IV solutions multiple times per day; use oral rehydration whenever possible; minimize or eliminate “keep vein open” (KVO) infusions, and decrease the use of flush bags for intermittent infusions. A revised (expanded) list of recommendations were sent on 10/10 specific to the children’s hospital, which included additions such as diligently optimizing diuresis to avoid over-diuresis, consider fluid boluses of albumin in hemodynamic instability over maintenance IVF, enteral electrolyte repletion, and commit to IV push medications whenever safe and possible. On 10/29/2024 the hospital updated its NPO time guidelines to align with ASA standards and help conserve additional fluid. These changes were implemented in order-specific decision support within the electronic health record.

Site 2: Notice of anticipated fluid restrictions was sent out via e-mail on 10/04/2024. Recommended strategies for conserving IVF included using enteral hydration whenever possible, waiting to spike IVF bag until needed, continuing use across patient care locations (ie, Emergency Department to floor, operating room to floor) and from outside hospitals where possible, and extending use of IVF bag from 4 to 7 days. Limiting IV flush volumes was also recommended by changing to enteral or intramuscular forms of medication administration where possible and giving IV medications via push or over syringe pump that required smaller flush volumes.

Site 3: On 11/1/2024, Site 3 had a meeting between head ICU pharmacist, nurse manager, and physician lead to come up with a single page IVF conservation plan. This was sent to the Chief Medical Officer, and then discussed at meetings with faculty and nursing on 11/5. Physician lead discussed with faculty easy changes to make including: converting IV medications to enteral (already in process at Site 3 as part of safety checklist), limiting/decreasing maintenance IVF (especially in the peri-operative setting), initiating enteral feeding earlier, and avoiding switching maintenance IVF type if not absolutely necessary. Pharmacy pushed use of bioavailable enteral medications and raised awareness of high-volume medications that could be concentrated. Nursing was asked to notify physician when IVF bags were nearly completion so the team could reassess necessity of another bag. They also made a push to avoid small volume infusions for vessel patency (“KVO”) and switch more to saline locking peripheral IVs.

Site 4: Shortly after Hurricane Helene Site 4 leadership sent out emails encouraging fluid restriction as clinically relevant. Shortly thereafter ICU teams received guidelines that recommended discontinuing unnecessary IVF (ex. when a patient is tolerating oral intake), using heparin locks, using saline flushes if small fluid volume was necessary, and changing IV medications to enteral when possible. Finally, we received more formal guidelines that included clinical actions to decrease use of IVF and parenteral nutrition as well as TPN inclusion/initiation criteria.
